# Prevalence of SARS-CoV-2 infection in previously undiagnosed health care workers at the onset of the U.S. COVID-19 epidemic

**DOI:** 10.1101/2020.04.20.20072470

**Authors:** Emily S. Barrett, Daniel B. Horton, Jason Roy, Maria Laura Gennaro, Andrew Brooks, Jay Tischfield, Patricia Greenberg, Tracy Andrews, Sugeet Jagpal, Nancy Reilly, Martin J. Blaser, Jeffrey L. Carson, Reynold A. Panettieri

**Affiliations:** Department of Biostatistics and Epidemiology; Rutgers School of Public Health; Piscataway, NJ; Environmental and Occupational Health Sciences Institute; Rutgers University; Piscataway, NJ; Department of Pediatrics, Rutgers Robert Wood Johnson Medical School; New Brunswick, NJ; Rutgers Center for Pharmacoepidemiology and Treatment Science, Institute for Health, Health Care Policy and Aging Research; New Brunswick, NJ; Public Health Research Institute; Department of Medicine; New Jersey Medical School; Rutgers University; Newark, NJ; RUCDR Infinite Biologics and Human Genetics Institute of NJ and Department of Genetics, Rutgers University; Piscataway, NJ; Department of Medicine, Rutgers Robert Wood Johnson Medical School; New Brunswick, NJ; Center for Advanced Biotechnology and Medicine; Rutgers University, Piscataway, NJ; Rutgers Institute for Translational Medicine & Science; New Brunswick, NJ

**Author notes:** joint first authorship (Emily S. Barrett, Daniel B. Horton). joint senior authorship (Martin J. Blaser, Jeffrey L. Carson, Reynold A. Panettieri). Corresponding author: Jeffrey Carson, MD, Division of General Internal Medicine, Rutgers, Robert Wood Johnson Medical School, 125 Paterson Street, New Brunswick, New Jersey 08901.

## Abstract

**Importance:** Healthcare workers are presumed to be at increased risk of severe acute respiratory syndrome coronavirus-2 (SARS-CoV-2) infection due to occupational exposure to infected patients. However, no epidemiological study has examined the prevalence of SARS-CoV-2 infection in a cohort of healthcare workers during the early phase of community transmission.

**Objective:** To determine the baseline prevalence of SARS-CoV-2 infection in a cohort of previously undiagnosed healthcare workers and a comparison group of non-healthcare workers.

**Design:** Prospective cohort study

**Setting:** A large U.S. university and two affiliated university hospitals

**Participants:** 546 health care workers and 283 non-health care workers with no known prior SARS-CoV-2 infection

**Exposure:** Healthcare worker status and role

**Main outcome(s) and measure(s):** SARS-CoV-2 infection status as determined by presence of SARS-CoV-2 RNA in oropharyngeal swabs.

**Results:** At baseline, 41 (5.0%) of participants tested positive for SARS-CoV-2 infection, of whom 14 (34.2%) reported symptoms. The prevalence of SARS-CoV-2 infection was higher among healthcare workers (7.3%) than in non-healthcare workers (0.4%), representing a 7.0% greater absolute risk (95% confidence interval for risk difference 4.7%, 9.3%). The majority of infected healthcare workers (62.5%) worked as nurses. Positive tests increased across the two weeks of cohort recruitment in line with rising confirmed cases in the hospitals and surrounding counties.

**Conclusions and relevance:** In a prospective cohort conducted in the early phases of community transmission, healthcare workers had a higher prevalence of SARS-CoV-2 infection than non-healthcare workers, attesting to the occupational hazards of caring for patients in this crisis. Baseline data reported here will enable us to monitor the spread of infection and examine risk factors for transmission among healthcare workers. These results will inform optimal strategies for protecting the healthcare workforce, their families, and their patients.

Clinicaltrials.gov registration number:NCT04336215

**Key points:** *Question:* Among previously undiagnosed individuals, is the prevalence of SARS-CoV-2 infection higher in U.S. healthcare workers compared to non-healthcare workers in the early phase of the U.S. COVID-19 epidemic?

*Findings:* The prevalence of SARS-CoV-2 infection was 7.3% in healthcare workers and 0.4% in non-healthcare workers, representing 7.0% greater absolute risk in the former (95% confidence interval for risk difference 4.7%, 9.3%). Infections were most common among nursing staff.

*Meaning:* Health care workers, particularly those with high levels of close patient contact, may be particularly vulnerable to SARS-CoV-2 infection. Additional strategies are needed to protect these critical frontline workers.

## Introduction

Healthcare workers (HCW) are a critical, yet understudied population during the current coronavirus disease-2019 (COVID-19) pandemic. On the frontlines of defense against the virus, HCW may experience increased risk of severe acute respiratory syndrome coronavirus-2 (SARS-CoV-2) infection due to close contact with highly infectious patients^1^ and, commonly, insufficient access to personal protective equipment (PPE)^2^. The plight of HCW during the pandemic has been widely noted^3-8^; as of April 9, 2020, 9,282 known COVID-19 cases among U.S. HCW had been reported to the Centers for Disease Control and Prevention (CDC) ^9^. Of HCW with COVID-19, 55% reported that their only known exposure was the in the health care setting.

However, our understanding of exposure among U.S. HCW is hindered by several key issues. First, there is clear underreporting of infection in this critical population as CDC data indicate that 84% of all reported U.S. COVID cases had no information on HCW status ^9^. Second, among both HCW and non-HCW (NHCW), access to testing has been inconsistent in the U.S., and a large proportion of cases that are asymptomatic or have only mild symptoms are likely to have gone untested ^10,11^. Importantly, asymptomatic or mildly symptomatic individuals can still transmit the virus and may represent the population most likely to spread the infection^12-14^. The rapid spread of the disease and high clinical demands on the HCW population during the pandemic have impaired efforts to prospectively and systematically study the prevalence of SARS-CoV-2 infection in U.S. HCWs. These data are vitally important to understand potential sources of exposure as well as to inform clinical decision-making about staffing and protections for HCW and their patients.

To this end, we report on the prevalence of SARS-CoV-2 infection in previously undiagnosed HCW and NHCW recruited into a prospective observational study conducted within a major U.S. academic medical system located in New Jersey, one of the U.S. epicenters of the pandemic.

## Methods

### Study population

From March 24-April 7, 2020, participants were recruited into the Rutgers Corona Cohort (RCC) at Rutgers University and participating university hospitals, University Hospital (Newark, NJ) and Robert Wood Johnson University Hospital (New Brunswick, NJ). Eligible HCW reported: (1) ≥20 hours of hospital work weekly; (2) occupations with regular patient exposure (e.g., residents, fellows, attending physicians, dentists, nurse practitioners, physician assistants, registered nurses, technicians, respiratory therapists, physical therapists); and (3) regular direct patient contact (≥3 patients/shift) expected in the next 3 months. Eligibility criteria for NHCW included: (1) faculty, staff, trainees, or students working at Rutgers ≥20 hours weekly; and (2) no patient contact. For both groups, additional eligibility criteria were: (1) ≥age 20; (2) not pregnant or breastfeeding; (3) no urgent care or emergency room visits, hospitalizations, operations, or changes in prescription medicines in the prior 30 days; and (4) no previously diagnosed SARS-CoV-2 infection or COVID-19.

### Study activities

An online pre-screener was used to determine eligibility. After informed consent, participants completed an online baseline questionnaire with items on demographics, general health, recent symptoms, lifestyle, occupation, and potential COVID-19 exposure followed by a face-to-face baseline visit. Trained study staff wearing personal protective equipment (PPE) measured body temperature and collected oropharyngeal swabs [OPS]. Participants presenting with fever ≥100.4°F were excluded. Study data were collected and managed using REDCap electronic data capture tools hosted at Rutgers Robert Wood Johnson Medical School^15^. All study activities were approved by the Rutgers Institutional Review Board prior to study implementation (Pro2020000679).

### SARS-CoV-2 assays

Assays were conducted under FDA approved EUA#200090. Following collection, Dacron OPS were immersed in phosphate-buffered saline (PBS) and transported at room temperature to RUCDR Infinite Biologics® (Piscataway, NJ) within 2 hours. Total RNA was extracted with Chemagic 360 (PerkinElmer) automation utilizing paramagnetic beads that bind nucleic acids (Chemagic Viral DNA/RNA 300 Kit H96). This system eliminates manual sample handling, reduces risk of cross-contamination and ensures rapid and consistent processing. Reverse transcriptase-PCR (RT-PCR) was performed using the Applied Biosystems TaqPath COVID-19 Combo Kit with 5μL of the extracted RNA sample. The Rutgers Clinical Genomic Laboratory TaqPath SARS-CoV-2 assay targets by quantitative real-time reverse transcription-PCR (RT-qPCR) three specific genomic regions of the SARS-CoV-2 genome; the nucleocapsid (N) gene, spike protein (S) gene, and ORF1ab region. There are positive and negative assay controls and MS2 phage is a positive control of nucleic acid extraction and RT-PCR. Assays were performed in triplicate. The lower limit of SARS-CoV-2 detection is 200 copies/mL and the assay exhibits no cross-reactivity with 43 organisms and viruses tested.

Participants who tested positive for SARS-CoV-2 were informed by a study physician, who assessed participants’ clinical condition and provided guidance on medical care, self-isolation, and cleaning ^16^.

### Statistical analysis

We evaluated and compared characteristics of HCW and NHCW using frequencies and chi-square testing. Confidence intervals were produced using 10,000 nonparametric bootstrap samples. All analyses were performed using SAS 9.4.

## Results

The cohort of 829 subjects [546 HCW and 283 NHCW] was predominantly female (63.9%), and half (51.6%) were <40 years old (Table 1). One-third (34.9%) of participants reported having at least 1 chronic medical condition and 4.5% reported currently smoking. Small proportions of HCW and NHCW (12.6% and 7.1%, respectively) reported contact with individuals outside of work with COVID-19 or suggestive symptoms (Table 1).

**Table 1.** Characteristics of the Rutgers Corona Cohort.

|  | **Total cohort**<br>**(n= 829)** | **HCW**<br>**(n=546; 65.9%)** | **NHCW**<br>**(n=283; 34.1%)** | **p-value<sup>1</sup>** |
| --- | --- | --- | --- | --- |
| **Demographics** |  |  |  |  |
| Female | 530 (63.9%) | 354 (64.8%) | 176 (62.2%) | 0.30 |
| Age (years) |  |  |  | 0.001 |
| 20-39 | 428 (51.6%) | 300 (55.0%) | 128 (45.2%) |  |
| 40-59 | 315 (38.0%) | 204 (37.4%) | 111 (39.2%) |  |
| ≥60 | 86 (10.4%) | 42 (7.7%) | 44 (15.6%) |  |
| Race |  |  |  | <0.001 |
| White | 483 (58.3%) | 285 (52.2%) | 198 (70.0%) |  |
| Asian | 168 (20.3%) | 129 (23.6%) | 39 (13.8%) |  |
| Black | 90 (10.9%) | 70 (12.8%) | 20 (7.1%) |  |
| Other/missing | 88 (10.6%) | 62 (11.4%) | 26 (9.2%) |  |
| Hispanic ethnicity | 100 (12.1%) | 67 (12.3%) | 33 (11.7%) | 0.80 |
| Current smoker | 37 (4.5%) | 19 (3.5%) | 18 (6.4%) | 0.06 |
| **Clinical characteristics** |  |  |  |  |
| Any chronic comorbidity<sup>2</sup> | 289 (34.9%) | 185 (33.9%) | 104 (36.8%) | 0.41 |
| Diabetes mellitus | 48 (5.9%) | 34 (6.3%) | 14 (5.0%) | 0.43 |
| Hypertension | 125 (15.2%) | 80 (14.9%) | 45 (15.9%) | 0.70 |
| Coronary or cerebrovascular disease | 20 (2.4%) | 15 (2.8%) | 5 (1.8%) | 0.37 |
| Asthma, COPD, or other chronic lung disease | 114 (13.9%) | 74 (13.8%) | 40 (14.2%) | 0.87 |
| Autoimmune disease or reported immunosuppressant use | 40 (4.9%) | 28 (5.2%) | 12 (4.2%) | 0.54 |
| COVID-19 symptoms in last week (any)<sup>3</sup> | 98 (11.9%) | 75 (13.9%) | 23 (8.1%) | 0.02 |
| Recent exposure outside of work to someone with COVID-19 or new fever, cough, or shortness of breath | 88 (10.7%) | 68 (12.6%) | 20 (7.1%) | 0.02 |

COVID-19: coronavirus disease-2019; HCW: healthcare workers; NHCW: non-healthcare workers
1 Chi-squared tests
2 Chronic comorbidities included diabetes mellitus, hypertension, coronary or cerebrovascular disease, heart failure, asthma, chronic obstructive pulmonary disease, other chronic lung disease, or chronic autoimmune disease
3 COVID-19 symptoms included fever, cough, shortness of breath, vomiting, diarrhea, or change in smell or taste.

Overall, 40 HCW (7.3%) and 1 NHCW (0.4%) tested positive for SARS-CoV-2 infection, representing 7.0% greater absolute risk (95% confidence interval for risk difference 4.7%, 9.3%) of SARS-CoV-2 among HCW compared to NHCW. The majority (65.9%) of infected participants reported no symptoms of infection in the previous week and no close contact with individuals outside of work who had symptoms or diagnoses of COVID-19 (82.9%). Among HCW, 71% reported working with at least one patient per shift who was known or suspected to be COVID-19 positive. Nurses had the highest rate of observed infection (11.1% positive) compared to 1.8% of attending physicians and 3.1% of resident and fellow physicians (Table 2). On average, nurses reported spending median 50% (interquartile range [IQE]: 40, 100%) of their time in patient rooms with a median of 10 (IQR: 6, 20) patient contacts per shift, during which they used PPE in a median of 100% (IQR 50, 100%) of patient contacts. By contrast, other HCW roles typically reported working a median of 20% (IQR 10, 50%) time in patients’ rooms with a median 10 (IQR 5, 15) patient contacts per shift, during which they used PPE in a median of 80% (IQR 30, 100%) of patients. ICU workers had low rates of observed infection (2.1%) compared to those working on other units (4.9-9.7%).

**Table 2.** Rates of SARS-CoV-2-infection among healthcare workers at two New Jersey hospitals (Robert Wood Johnson University Hospital [RWJUH] and University Hospital Newark [UHN]).

| Variable | # SARS-CoV-2 + /<br>total n (%)<br>40/546 (7.3%) | # SARS-CoV-2 + /<br>n at RWJUH (%)<br>10/291 (3.4%) | # SARS-CoV-2 + /<br>n at UHN (%)<br>30/255 (11.8%) |
| --- | --- | --- | --- |
| **Health care role** |  |  |  |
| Attending physician | 2/112 (1.8%) | 1/63 (1.6%) | 1/49 (2.0%) |
| Resident or fellow physician | 3/98 (3.1%) | 2/63 (3.2%) | 1/35 (2.9%) |
| Nurse | 25/225 (11.1%) | 7/123 (5.7%) | 18/102 (17.7%) |
| Other | 10/111 (9.0%) | 0/42 (0%) | 10/69 (14.5%) |
| **Primary unit<sup>1</sup>** |  |  |  |
| Emergency department | 20/242 (8.3%) | 8/130 (6.2%) | 12/112 (10.7%) |
| Medical floor | 9/184 (4.9%) | 5/103 (4.9%) | 4/81 (4.9%) |
| Operating room | 13/134 (9.7%) | 1/53 (1.9%) | 12/81 (14.8%) |
| Intensive care unit<sup>2</sup> | 4/192 (2.1%) | 1/111 (0.9%) | 3/81 (3.7%) |
| Other unit | 17/280 (6.1%) | 4/138 (2.9%) | 13/142 (9.2%) |
| **Estimated percentage of work-time spent in patients' rooms** |  |  |  |
| <25% | 11/210 (5.2%) | 1/116 (0.9%) | 10/94 (10.6%) |
| 25-49% | 7/114 (6.1%) | 1/62 (1.6%) | 6/52 (11.5%) |
| 50-74% | 11/116 (9.5%) | 6/66 (9.1%) | 5/50 (10%) |
| ≥75% | 11/95 (11.6%) | 2/39 (5.1%) | 9/56 (16.1%) |
| Missing | 0/11 (0%) | 0/8 (0%) | 0/3 (0%) |
| **Estimated percentage of patients for which PPE<sup>3</sup> was used** |  |  |  |
| <25% | 4/87 (4.6%) | 2/53 (3.8%) | 2/34 (5.9%) |
| 25-49% | 2/58 (3.5%) | 1/31 (3.2%) | 1/27 (3.7%) |
| 50-75% | 4/60 (6.7%) | 1/29 (3.5%) | 3/31 (9.7%) |
| 75-99% | 4/41 (9.8%) | 2/17 (11.8%) | 2/24 (8.3%) |
| 100% | 25/269 (9.3%) | 4/147 (2.7%) | 21/101 (17.2%) |
| Missing | 1/31 (3.2%) | 0/14 (0%) | 1/17 (5.9%) |
| **Average number of patients with suspected or confirmed COVID-19 per shift<sup>4</sup>** |  |  |  |
| 0 | 7/158 (4.4%) | 2/89 (2.3%) | 5/69 (7.3%) |
| >0-<5 | 9/162 (5.6%) | 0/89 (0%) | 9/73 (12.3%) |
| ≥5 | 24/226 (10.6%) | 8/113 (7.1%) | 16/113 (14.2%) |
| Missing | - | - | - |

COVID-19 coronavirus disease-2019; PPE personal protective equipment; RWJUH Robert Wood Johnson University Hospital; SARS-CoV-2 severe acute respiratory syndrome coronavirus-2; UHN University Hospital Newark
1 Half (50.4%) of HCW, including 15 (37.5%) of those infected, reported more than one primary unit.
2 Included medical, surgical, cardiac, neurocritical, pediatric, and neonatal intensive care units.
3 Personal protective equipment referred to wearing gloves, gown, and a mask (surgical or N95)
4 Decimal places allowed in response

Half of the HCW reported using PPE during all patient contacts, but only 9.3% of those reporting universal PPE use tested positive. Rates of SARS-CoV-2 infection were slightly higher among workers who spent greater proportions of time in patients’ rooms, reported higher levels of PPE use, and reported exposure to more patients with suspected or diagnosed COVID-19 (Table 2).

During the two weeks of participant recruitment, the daily frequency of SARS-CoV-2 positivity rose among HCW and was consistent with increases in confirmed infections among both participating hospitals as well as the surrounding counties (Figure 1). During the enrollment period, higher rates of SARS-CoV-2 infections were seen at University Hospital Newark, which had a higher proportion of COVID-19 patients and is located in a geographic area with higher infection rates (Figure 1, Table 2).

**Figure 1.**
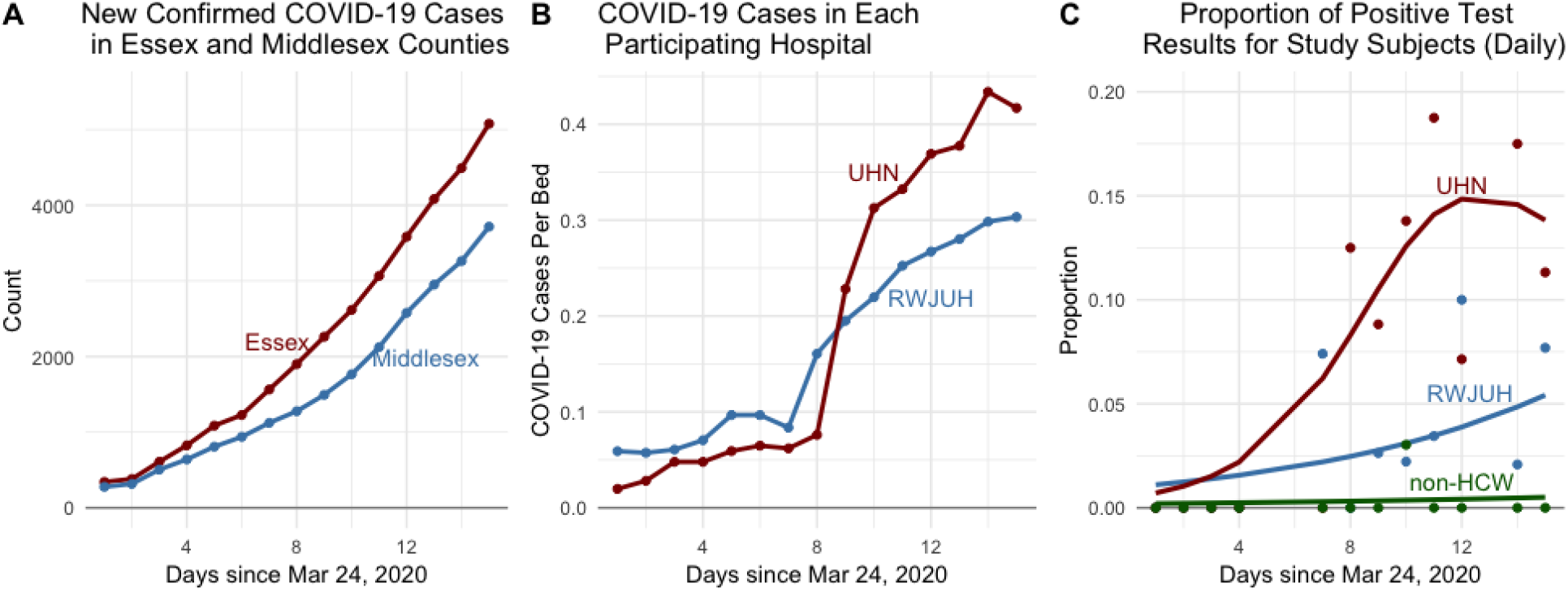
Prevalence of SARS-CoV-2 infection in a prospective cohort, participating hospitals, and surrounding counties during the study period (3/24/2020-4/7/2020). (A) confirmed cases of COVID-19 in New Jersey counties containing the participating hospitals [Essex County – University Hospital Newark; Middlesex County-Robert Wood Johnson University Hospital]; (B) confirmed inpatient cases of COVID-19 per total hospital beds in participating hospitals; and (C) confirmed SARS-CoV-2 positive cases in healthcare workers (HCW) and non-healthcare workers (NHCW) by hospital in the Rutgers Corona Cohort. County data comes from the New Jersey Department of Health (as reported in the New York Times). RWJUH Robert Wood Johnson University Hospital; SARS-CoV-2 severe acute respiratory syndrome coronavirus-2; UHN University Hospital Newark

## Discussion

In a prospective cohort of 829 individuals without previous diagnoses of SARS-CoV-2 infection or COVID-19, 7.3% of HCW and 0.4% of NHCW were found to be SARS-CoV-2-positive. These results support the hypothesis of higher SARS-CoV-2 prevalence in HCW compared with NHCW, a difference which may be attributable to workplace exposures, given the low rate of infection in NHCW. While HCW were slightly more likely than NHCW to report sick contacts outside of work, fewer than 1 in 5 HCW reported having a sick contact. SARS-CoV-2-infected HCW appeared more likely to be nurses, to spend more time in patients’ rooms, and to have more patients with suspected or confirmed COVID-19. We also observed a lower prevalence of SARS-CoV-2 among ICU workers compared to other units. HCW in our cohort who reported lower usage of PPE did not appear to have higher rates of infection, suggesting that use of protective measures were proportional to their perceived risk of acquiring infection.

Over the two-week recruitment period, there was an apparent increase in the number of participating HCW (but not NHCW) testing positive for SARS-CoV-2. This rise was consistent with the sharp increase in confirmed number of positive cases in the participating hospitals and well as the surrounding areas ^17^. With its proximity to New York City (NYC), NJ is one of the states hardest hit by COVID-19 crisis to date: home to less than 3% of the U.S. population^18^, it has over 75,000 confirmed COVID-19 cases, representing 11.7% of all known cases nationwide ^17,19^. The first confirmed COVID positive case in NJ was reported on March 3, 2020. On March 9, the governor declared a state of emergency followed soon after by a statewide curfew (March 16) and stay-at-home order (March 21). At the time our recruitment began on March 24, 2020, 3,675 cases had been reported in NJ, second only to New York; as of the time of writing (April 17), there are over 75,000 confirmed cases in the state indicating the ongoing magnitude of the crisis^17^. Compared to Robert Wood Johnson University Hospital situated in Central NJ (Middlesex County), the infection rate among HCW was nearly 3.5 times higher at University Hospital Newark, an urban hospital situated close to NYC (Essex County) with higher population density and higher rates of infections, as well as a higher proportion of hospitalized patients with COVID-19. The difference in infection rates detected among HCW at these two hospitals highlights the variability across hospitals even within the same medical system and the need for more research on COVID-19 in HCW across diverse healthcare settings.

Several studies have reported on the high burden of infections among HCW, including cross-sectional and retrospective studies of symptomatic or hospitalized HCW^3,5,7,13^. By contrast, our findings derive from a prospective cohort of HCW without known SARS-CoV-2 infection or COVID-19 diagnosis at the time of screening. Our study also represents the first prospective study directly comparing rates of SARS-CoV-2 infection between HCW and NHCW.

Limitations of this study include opportunistic recruitment that may have led to over-enrollment of subjects highly concerned about potential infection. In our cohort, slightly higher proportions of HCW versus NHCW reported recent COVID-19 symptoms or sick contacts with COVID-19 diagnoses or symptoms, raising the possibility of ascertainment bias. However, simultaneous enrollment and testing of NHCW in the same locations and timeline, and the low overall prevalence of recent symptoms or exposures among both HCW and NHCW minimized this source of bias. Despite biases that could have raised the rates of detected infections, in fact 95% of the cohort were uninfected. We cannot definitively identify the exposures leading to infection or rule out HCW infections transmitted from sources other than hospitalized patients, including asymptomatic colleagues or contacts outside the hospital ^13^. The hospital with higher rates of infected HCW had both higher rates of infected patients within the facility as well as higher rates of infections in the surrounding area. However, infected HCW appeared to spend more time in patients’ rooms and care for more patients with COVID-19, and HCW were considerably more likely than NHCW to be infected, suggesting possible infectious transmission within the hospital. Planned longitudinal follow-up in this cohort will provide novel incidence and exposure data. Finally, the small numbers of SARS-CoV-2 diagnoses limited statistical comparisons.

In summary, in a prospective cohort of individuals previously undiagnosed with SARS-CoV-2, conducted in the early phases of community transmission, HCW had a considerably higher prevalence of SARS-CoV-2 infection than NHCW. We observed higher rates of infection among nurses, in those caring for more patients with suspected or confirmed COVID-19, and in the hospital with a higher proportion of patients with COVID-19. Lower rates of PPE use did not appear to correspond to higher rates of infection. Most infected participants reported no symptoms of COVID-19 and had no known sick contacts outside of the workplace. Additional strategies are needed to protect these critical frontline workers and to identify deficiencies in current protections. This prospective HCW cohort provides the baseline data to study the incidence and other risk factors of SARS-CoV-2 infection in this crucial population as the pandemic advances.

## Data Availability

Data may be made available to potential collaborators once the primary analyses have been completed by the study team.

## Acknowledgments

We would like to acknowledge the tireless work of the entire Rutgers Corona Cohort study team, all based at Rutgers University: for conducting study visits, we thank Kathleen Black, PhD, MPH, Deborah McCloskey, RN, BSN, Taylor Black, MPH, Adriana De Resende, Alicia Legard, Christie Lyn Costanza, MPH, Christina Dalliani, Eric Asencio, Jared Khan, MPH, Valorie Cadorett, Ariana Alcaide, MPH, Kathleen Bott, RN, Randall Teeter, Juan Diego Ramirez, Adriana Hemans, Eliana Obando-Jaramillo, Yanille Taveras, MS, Marisol Rivera, Lisa Cerracchio, RNC, Halina Malveaux, RN, Jessica Kirby-Smith, Fei Chen, RN, Christina Varghese, William Russell, RPFT, Marie Macor, RN, Anthony Helena, Karen Dragert, RN, Aura Velasco, LPN, Susette Coyle, RN, MSN, Ami Patel, and Elizabeth George, RPH, BCOP; for providing supplies, we thank Joseph Barone, PharmD, and Daniel Notterman, MD; for facilitating implementation of the study at the participating clinical sites, we thank: Brian Buckley PhD, Debra Chew, MD, MPH, Mark Einstein, MD, MS, Shereef Elnahal, MD, Cecile Feldman, DMD, MBA, John Gantner, MBA, CPA, Sununda Gaur, MD, Brian Strom, MD, MPH, Stanley Z. Trooskin, MD, and Helmut Zarbl, DCS, PhD,; for assistance with NJ state data, we thank Panos Georgopoulos, PhD, Zhongyuan Mi, MPH, and Xiang Ren, MS; for their administrative assistance, we thank: Helaine Novek, Judith Argon, MS, Veenat Parmar, Matthew Leonardelli; for assistance with literature review, we thank Michael Hoven.

In-kind logistics support was provided by Marken/UPS and dfYoung.

Finally, we thank our brave participants serving on the frontlines of the pandemic. The authors report no conflicts of interest.

